# Respiratory rates among rural Gambian children: a community-based cohort study

**DOI:** 10.1101/2023.12.05.23299490

**Authors:** Polycarp Mogeni, Sharon Amima, Jennifer Gunther, Margaret Pinder, Lucy S. Tusting, Umberto D’Alessandro, Simon Cousens, Steve W. Lindsay, John Bradley

**Affiliations:** Department of Infectious Disease Epidemiology, London School of Hygiene & Tropical Medicine, London, UK; Kenya Medical Research Institute (KEMRI), Nairobi, Kenya; Department of Food Science, Nutrition and Technology, University of Nairobi, Nairobi, Kenya; Department of Biosciences, Durham University, Durham, UK; Medical Research Council’s (MRC) Unit The Gambia at the London School of Hygiene and Tropical Medicine, Banjul, The Gambia; Department of Disease Control, London School of Hygiene & Tropical Medicine, London, UK

## Abstract

**Background:** Although ranges of normal respiratory rates (RR) have been described for children under five years old living in the tropics, there are few datasets recording rates in older children. The present study was designed to capture the changes in RR with age and to examine its association with nutritional status and environmental factors.

**Methods:** A cohort of rural Gambian children aged from six months to 14 years had their RR recorded during home visits twice weekly during two annual rainy seasons. Measurements were made by trained field assistants using an electronic timer during a one-minute period. Age, sex, nutritional status, health status, time of day of data collection were recorded. A generalized additive model for location, scale and shape was used to construct the RR reference curves and a linear mixed effect model used to examine factors associated with RR. We also assessed the agreement between repeat measurements taken from a subset of study subject.

**Results:** A total of 830 children provided 67,512 RR measurements. The median age was 6.07 years (interquartile range (IQR), 4.21–8.55) and 400 (48.2%) were female. The centile chart showed a marked nonlinear decline in RR measurements with increasing age up to six years old, after which the decline was minimal (predicted median RR of 31 breaths/minute (IQR: 29–34) among one-year-olds, 22 breaths/minute (IQR: 21–23) among six-year-olds and 21 breaths/minute (IQR: 21-22) among 13-year-olds. Age (non-linear effect, p<0.001), stunting (0.84 breaths/minute [95%CI: 0.40-1.28, p<0.001]), ambient temperature (0.38 breaths/minute [95%CI: 0.33-0.42, p<0.001] for every 1°C increase in ambient temperature) and time of day when RR measurements were taken (non-linear effect, p<0.001) were independent predictors of respiratory rate. Strikingly, children with signs of illness were associated with higher intra-observer variability.

**Interpretation:** We constructed a RR reference chart for children aged one to 13 years and proposed a cutoff of >26 breaths/minute for raised RR among children aged >5 years bridging an important gap in this age group. Although time of data collection, nutritional status and ambient temperature were predictors of RR, the evidence is not clinically significant to warrant a change in the current WHO guidelines owing to the prevailing uncertainty in the measurement of RR. The finding that RR between repeat measurements were more variable among children with signs of illness suggests that a single RR measurements may be inadequate to reliably assess the status of sick children - a population in which accurate diagnosis is essential to enable targeted interventions with lifesaving treatment.

## Background

Childhood respiratory illness constitutes the largest cause of post neonatal mortality globally, a burden that could be considerably reduced by timely diagnosis and treatment (1–3). Respiratory rate (RR) is an important vital sign monitored in clinical settings for clinical deterioration and respiratory distress, and is used to support the diagnosis of severe respiratory infections (4–6). As an indicator of respiratory function, healthcare professionals use RR to provide timely intervention, manage disease, and monitor treatment effect (6).

The World Health Organization (WHO) defines tachypnoea (abnormally rapid breathing) as a RR ≥60 breaths per minute among children aged <2 months, RR ≥50 among children aged between 2 to <12 months, and RR ≥40 among children aged 12 months to <60 months (4). However, RR measurement in children is often challenging (7,8) and currently there are no guidelines for children older than 5 years. RRs are typically counted manually in low-resource settings as breaths per minute using electronic timers and counting beads (9). Though manual measurement is often the gold standard in rural sub-Saharan Africa (sSA), it can be imprecise and subject to intra-observer variability compounded by whether the child is asleep or awake at the time when measurements are taken (8,10). In response to calls for better pneumonia diagnosis (11,12), automated RR counting aids are increasingly under development and evaluation (13–16). However, there are concerns that some of these aids are out of reach or are unsuitable in resource limited settings (8).

RR measurements are subject to variations associated with both intrinsic factors (e.g. child age, gender, underlying health conditions, child state (asleep, awake or agitated), underlying health conditions) and extrinsic factors that include environmental and socioeconomic factors (8,17). Younger children have higher and more variable RR measurements compared to older children whilst those residing at higher altitudes have higher RR measurements, likely due to lower oxygen concentration at higher altitudes (17–19).

Here, we report the results of 92 consecutive rounds of community follow-up of children recruited to participate in a trial investigating the effect of improved housing on the incidence of clinical malaria in The Gambia (20). The study was undertaken over two consecutive malaria transmission seasons between June and December of 2016 and 2017 (20). We utilized a large longitudinal dataset of RR measurements taken twice weekly to construct the RR-for-age reference curves for children aged 1-13 years of age and provide a comprehensive description of the predictors of RR among children residing in rural Gambia; a country at low altitude. This is one of the few studies to report RR in children aged >5 years and the first to do so among rural Gambian children.

## Methods

### Study setting

The study was conducted in villages in the Upper River Region of The Gambia, located in the east of the country. This is an area of Sudanian savanna at an altitude of less than 50 m above sea level. The mean annual rainfall is 876 mm which falls mainly between May and October, followed by a long dry season. The enrolled villages are located on both the north and south banks of the River Gambia; an area that experiences an average maximum daily temperature of 36°C and a minimum of 18°C. Further details have been reported previously (21).

### Study design and participants

Data came from a household randomized controlled trial involving children aged 6 months to 14 years, who were recruited to a study investigating the effect of improved housing on the incidence of clinical malaria (20,21). The study took place over a two-year period in which clinical follow-ups were conducted over two consecutive malaria transmission seasons (June to December of 2016 and 2017) (21). Details of the study design, data collection, consenting, supervision and training procedures have been presented previously in a protocol (21) and in the main study results (20). Briefly, alongside the primary trial endpoint of clinical malaria, respiratory rate data collection was done twice weekly on every participating child by trained health care providers equipped with electronic timers. The data were collected because of concerns that the housing intervention could cause respiratory illness due to restricting airflow through the house. The final trial results, however, showed no evidence of an increase in respiratory illness in the intervention group (20). Assessment was done twice weekly (21) to increase case detection rate given the high burden of childhood pneumonia in west Africa (22). The process involved the development of a standardized protocol containing guidelines for collecting all clinical measurements which included recording respiratory rate measurements when the child was awake and calm. If any measurement was ≥50 breaths/minute for children under 1 year, ≥40 breaths/minute for children aged 1 to 5 years or ≥25 breaths/minute among children aged 5 to 14 years of age, a repeat measurement was taken at least 5 minute after the initial measurement by the same staff member (20). Measurements were taken by observing the child’s naked chest movement, where one rise and fall of the chest was counted as one breath and recorded as the number of breaths per minute.

### Statistical analysis

#### Construction of reference curves

The main analytical objective was to construct a RR reference chart that can be used to inform clinical management of respiratory illnesses in low altitude areas. The outcome of interest was the RR measurement derived from children who were not reported to be sick during data collection in the household. Here, we excluded data from children who were unwell, defined by signs of respiratory illness (cough, chest indrawing, wheeze or difficulty in breathing), fever (axillary temperature ≥37.5°C), signs of gastrointestinal illness (vomiting and diarrhea), child reported as being unwell by caregiver or on medication (Figure 1). In addition, we excluded one outlier value (recorded as 181 breaths/minute) that is not plausible for the child’s age and because the measurement was not accompanied by a second measurement as per the requirements of the protocol (21).

**Figure 1.**
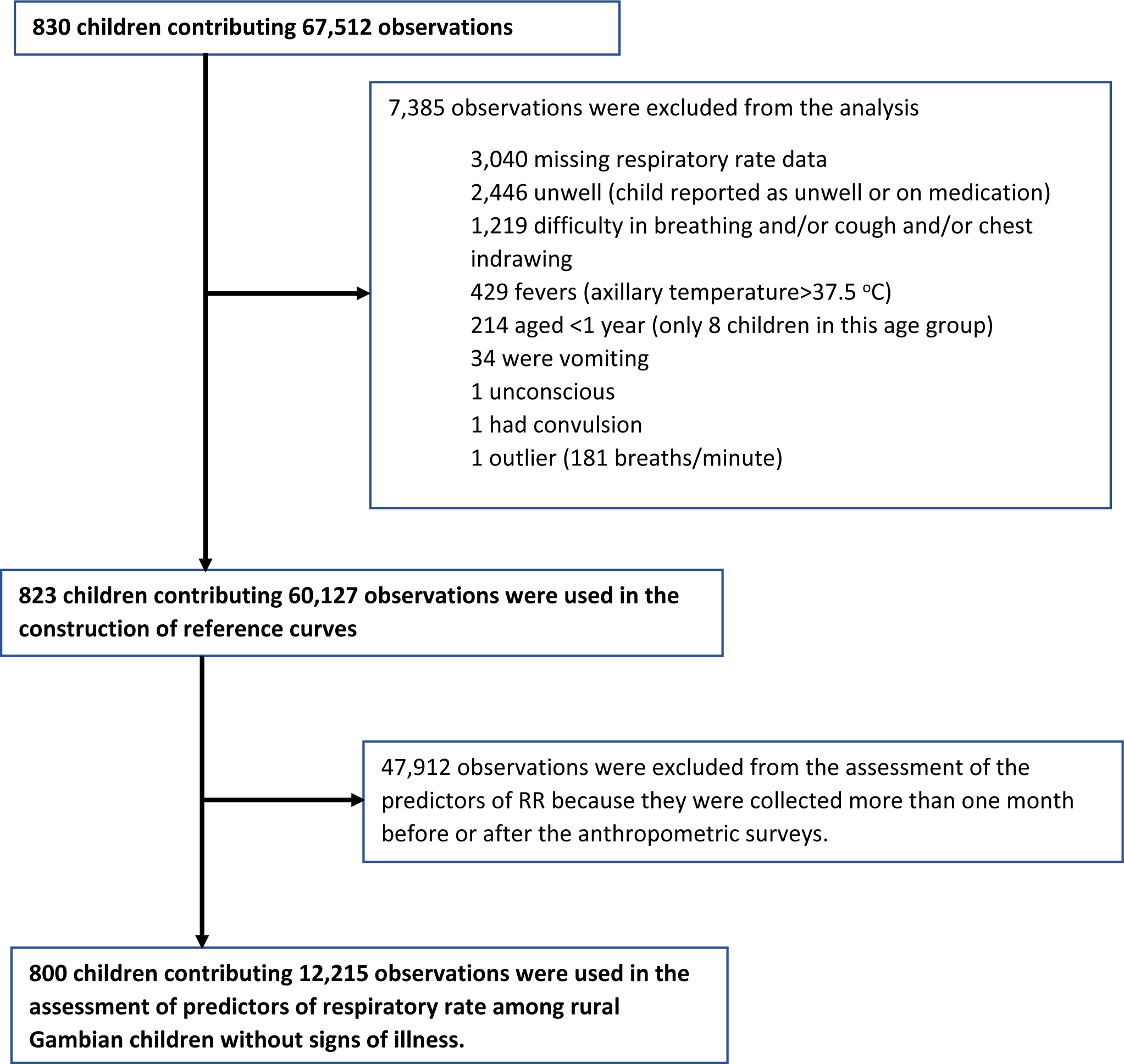
Number of participants, observations contributed and reasons for exclusion at each analysis stage.

We conducted an exploratory analysis to examine potential outliers in the dataset and visually inspected the association between respiratory rate and age using the lowess function and scatter plots. Smooth percentile curves of respiratory rate as a function of age were constructed using the generalized additive models for location, scale and shape (GAMLSS) (23). The Box-Cox-t model (24) was the best fit to our data among the plausible GAMLSS models (the Box-Cox-t, Box-Cox-Power-Exponential and Box-Cox-Cole-green) that were examined in a data driven model selection procedure (25,26). The selected model was used to construct the reference chart (at - 2 standard deviation (SD), −1SD, median, 1SD and 2SD) of RR as a function of age. The results were truncated at 1 and 13 years to reduce edge effects and the impact of the limited number of study participants <1 (n=8) and >13 years (n=32) of age.

#### Predictors of respiratory rate

The second analytical objective was to examine the predictors of respiratory rate with the aim of identifying potential subgroups that may influence the accuracy of the constructed reference chart. We considered nutritional status, rainfall, minimum and maximum daily temperature as a proxy for ambient temperature, time of day when the measurement was taken, sex and the randomization arm as potential predictors in the regression models. We linked the respiratory rate data to three cross-sectional anthropometric surveys conducted in December 2016, June 2017 and December 2017. The survey datasets were linked to the child’s corresponding respiratory rate observations that were collected within a month of each survey to reduce potential bias resulting from linking observations that are far apart in time. Anthropometric z-scores were calculated using the 2006 and 2007 WHO growth references for children age <5-years (25) and children aged ≥5-years (26) respectively. We defined stunting as a height-for-age z-score (HAZ) <-2, underweight as weight-for-age z-score (WAZ) <-2 and wasting as weight-for-height/length z-score (WHZ) <-2. Daily rainfall, minimum and maximum temperatures were obtained from the meteorological department in Basse Santa Su town.

Associations between respiratory rate and potential predictor variables were assessed using mixed-effects linear regression with random effects for each child. Age was categorized following the trend observed in the lowess curve to accommodate the non-linear trend. The likelihood ratio test statistic was used to compare a model with random intercepts only and the model with both random intercepts and slopes. Effect estimates and their corresponding 95% confidence intervals (95% CI) were used to assess the magnitude and direction of effect for the various predictor variables included in the regression models.

In an ancillary analysis, we assessed repeatability of RR measurements by examining the degree of agreement between repeat measurements that were taken within 5 minutes of each other using the Bland-Altman method of estimating the degree of agreement (28). These repeat measurements were taken only when the first respiratory rate measurement was deemed raised for the child’s age (20). We also examined the predictors of the absolute difference between paired repeat measurements using the linear regression model. Intra-observer variability was defined as the variation in repeated measurements by the same observer and used the intra-class correlation coefficient (ICC) to assess reliability of repeat measurements. The GAMLSS package (23) in R statistical environment was used to estimate the centile curves whilst the remaining analyses were conducted using Stata 17 (Stata Corp, College Station, TX, USA).

## Results

### Reference curves

A total of 830 children recruited to the study contributed 67,512 observations in the analyses. The median age was 6.07 years (interquartile range (IQR), 4.21 – 8.55) and 400 (48.2) were female. Details of study participants and the number of observations contributed at each stage of the analysis are presented in Figure 1. The RR for age reference chart was constructed using 60,127 observations from 823 children (Figure 2). There was a marked nonlinear decline in RR with increasing age between 1 and 6 years of age, after which the decline was minimal (the predicted median RR was 31 breaths/minute [IQR: 29–34] among 1 year-olds, 22 breaths/minute [IQR: 21–23] among 6 year-olds and 21 breaths/minute [IQR: 21-22] among 13 year-olds). The pre-specified cutoff for raised RR (>+2SD) declined from 46 breaths/minute among 1 year-olds to 27 breaths/minute among 6 year-olds and to 25 breaths/minute among 13 year-olds (Table 1, Figure 2).

**Figure 2:**
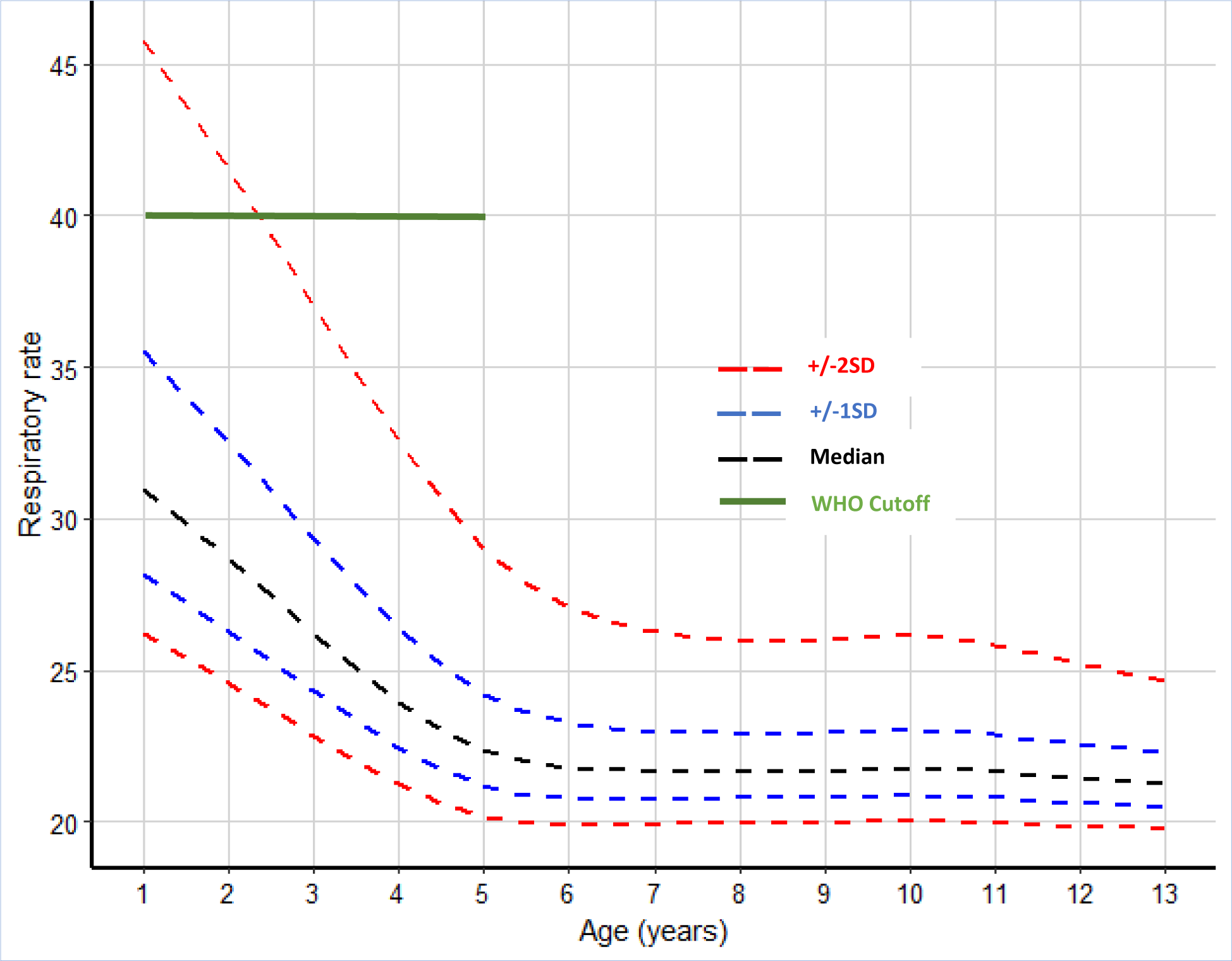
Respiratory rate (breaths/minute) for age reference curves. The black dashed line represents the median RR measurements, the blue dashed lines represent the 1SD from the median, the red dashed lines represent 2SD from the median and the green continuous line represents the WHO cutoff for tachypnoea (40 breaths/minute) among children aged >12 months and <5 years. Note that the WHO does not provide guidelines on tachypnoea among children aged above 5 years.

**Table 1:** Summary of children without signs of illness and their corresponding crude and model estimate of 2SD cutoff for raised respiratory rate (RR).

| Crude estimate of respiratory rate cutoff |  |  |  |  | GAMLSS estimate of RR cutoff |  |  |
| --- | --- | --- | --- | --- | --- | --- | --- |
| Age in Years | Observations | Number of Children | Median (IQR) | 2SD (Cutoff for raised RR) | Age in Years | Median (IQR) | 2SD (Model cutoff for raised RR) |
| 1 to <1.5 | 410 | 20 | 32 (28 - 35) | 41 | 1 | 31 (29 - 34) | 46 |
| 1.5 to <2.5 | 2609 | 89 | 32 (28 - 35) | 40 | 2 | 29 (27 - 31) | 42 |
| 2.5 to <3.5 | 4308 | 142 | 30 (26 - 34) | 38 | 3 | 26 (25 - 28) | 37 |
| 3.5 to <4.5 | 5273 | 177 | 28 (24 - 33) | 38 | 4 | 24 (23 - 25) | 33 |
| 4.5 to <5.5 | 7180 | 218 | 25 (21 - 30) | 38 | 5 | 22 (22 - 23) | 29 |
| 5.5 to <6.5 | 7875 | 237 | 21 (19 - 25) | 30 | 6 | 22 (21 - 23) | 27 |
| 6.5 to <7.5 | 6753 | 209 | 20 (19 - 24) | 29 | 7 | 22 (21 - 22) | 26 |
| 7.5 to <8.5 | 6593 | 193 | 20 (18 - 23) | 28 | 8 | 22 (21 - 22) | 26 |
| 8.5 to <9.5 | 5821 | 167 | 20 (18 - 24) | 28 | 9 | 22 (21 - 22) | 26 |
| 9.5 to <10.5 | 4600 | 142 | 19 (18 - 22) | 29 | 10 | 22 (21 - 22) | 26 |
| 10.5 to <11.5 | 3881 | 118 | 20 (18 - 22) | 28 | 11 | 22 (21 - 22) | 26 |
| 11.5 to <12.5 | 3092 | 92 | 19 (18 - 22) | 28 | 12 | 21 (21 - 22) | 25 |
| 12.5 to <13.5 | 1478 | 49 | 19 (17 - 22) | 26 | 13 | 21 (21 - 22) | 25 |

### Respiratory rate among sick children

A total of 4,119 (6.4%) observations from sick children were excluded from the construction of the reference chart. The median age of sick children was 5.5 years (IQR: 3.6–8.0) and 2016 (48.9%) observations were from female subjects. The distribution of observations from children with and without signs of illness by age and RR are presented in the Supplementary Table 1 and Supplementary Figure 1.

### Predictors of respiratory rate among community children without signs of illness

The prevalence of stunting (1 to 14 years of age) was 21.1% (95%CI: 19.4% to 22.9%), prevalence of underweight (1 to 10 years of age) was 22.6% (95%CI: 20.6% to 24.6%) and prevalence of wasting (1 to <5 years of age) was 8.0% (95%CI: 5.9% to 10.6%). Females contributed 1,037 (47.7%) observations in the 3 cross-sectional surveys. The distribution of observations over age and RR measurements are presented in Supplementary Figure 1A and 1C, and Table 1.

In a pooled adjusted mixed-effects linear regression (n=800 children, contributing 12,215 observations); age (non-linear effect), stunting, minimum daily temperature (a proxy for ambient temperature) and time of day of data collection were associated with respiratory rate (Table 2). On average, stunting was associated with an 0.84 breaths/minute (95%CI: 0.40-1.28, p<0.001) increase in RR. A 1°C increase in minimum daily temperature was associated with a 0.38 breaths/minute (95%CI: 0.33-0.42, p<0.001) increase in RR. In addition, there was a nonlinear relationship between RR rate measurements and the time of the day when the measurements were taken (Table 2). On average RR measurements increased from 0700hrs to ∼1500hrs after which the measurements declined towards 2000hrs (p<0.001). There was no evidence that being female or enrolled in the intervention group was associated with RR in the adjusted regression models (Table 2). As observed in the GAMLSS model and consistent with previous studies (18,19), older children had lower RR measurements in both unadjusted and the adjusted regression model (Table 2). That is, compared to children aged 1-to-2-year-old; children aged 2-to-3-years, 3-to-4 years, 4-to-6 years, 6-to-8 years and 8-to-13 years of age had RRs that were 0.38, 2.15, 5.14, 9.56 and 10.64 breaths/minute lower respectively (Table 2).

**Table 2:**
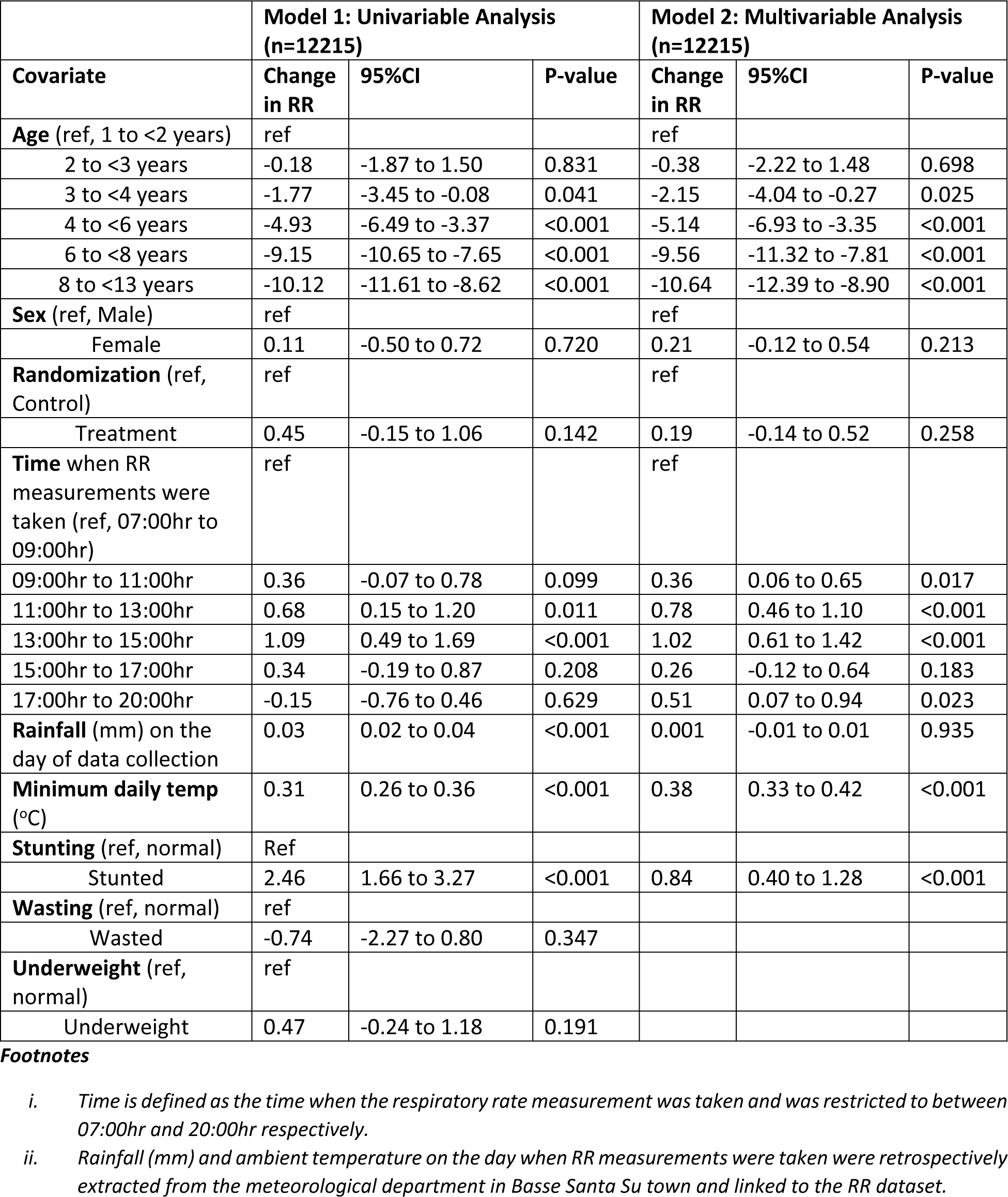
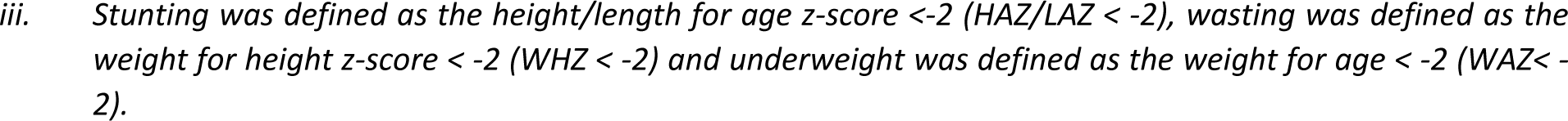
Predictors of respiratory rate. Model 1 presents results of the univariable mixed effect linear regression model and Model 2 presents results from the final multivariable mixed effect linear regression model. Sex and the randomization group have been included as potential confounders in the multivariable model.

|  | **Model 1: Univariable Analysis**<br>**(n=12215)** |  |  | **Model 2: Multivariable Analysis**<br>**(n=12215)** |  |  |
| --- | --- | --- | --- | --- | --- | --- |
| **Covariate** | **Change**<br>**in RR** | **95%CI** | **P-value** | **Change**<br>**in RR** | **95%CI** | **P-value** |
| **Age** (ref, 1 to <2 years) | ref |  |  | ref |  |  |
| 2 to <3 years | -0.18 | -1.87 to 1.50 | 0.831 | -0.38 | -2.22 to 1.48 | 0.698 |
| 3 to <4 years | -1.77 | -3.45 to -0.08 | 0.041 | -2.15 | -4.04 to -0.27 | 0.025 |
| 4 to <6 years | -4.93 | -6.49 to -3.37 | <0.001 | -5.14 | -6.93 to -3.35 | <0.001 |
| 6 to <8 years | -9.15 | -10.65 to -7.65 | <0.001 | -9.56 | -11.32 to -7.81 | <0.001 |
| 8 to <13 years | -10.12 | -11.61 to -8.62 | <0.001 | -10.64 | -12.39 to -8.90 | <0.001 |
| **Sex** (ref, Male) | ref |  |  | ref |  |  |
| Female | 0.11 | -0.50 to 0.72 | 0.720 | 0.21 | -0.12 to 0.54 | 0.213 |
| **Randomization** (ref, Control) | ref |  |  | ref |  |  |
| Treatment | 0.45 | -0.15 to 1.06 | 0.142 | 0.19 | -0.14 to 0.52 | 0.258 |
| **Time** when RR measurements were taken (ref, 07:00hr to 09:00hr) | ref |  |  | ref |  |  |
| 09:00hr to 11:00hr | 0.36 | -0.07 to 0.78 | 0.099 | 0.36 | 0.06 to 0.65 | 0.017 |
| 11:00hr to 13:00hr | 0.68 | 0.15 to 1.20 | 0.011 | 0.78 | 0.46 to 1.10 | <0.001 |
| 13:00hr to 15:00hr | 1.09 | 0.49 to 1.69 | <0.001 | 1.02 | 0.61 to 1.42 | <0.001 |
| 15:00hr to 17:00hr | 0.34 | -0.19 to 0.87 | 0.208 | 0.26 | -0.12 to 0.64 | 0.183 |
| 17:00hr to 20:00hr | -0.15 | -0.76 to 0.46 | 0.629 | 0.51 | 0.07 to 0.94 | 0.023 |
| **Rainfall** (mm) on the day of data collection | 0.03 | 0.02 to 0.04 | <0.001 | 0.001 | -0.01 to 0.01 | 0.935 |
| **Minimum daily temp** (°C) | 0.31 | 0.26 to 0.36 | <0.001 | 0.38 | 0.33 to 0.42 | <0.001 |
| **Stunting** (ref, normal) | Ref |  |  |  |  |  |
| Stunted | 2.46 | 1.66 to 3.27 | <0.001 | 0.84 | 0.40 to 1.28 | <0.001 |
| **Wasting** (ref, normal) | ref |  |  |  |  |  |
| Wasted | -0.74 | -2.27 to 0.80 | 0.347 |  |  |  |
| **Underweight** (ref, normal) | ref |  |  |  |  |  |
| Underweight | 0.47 | -0.24 to 1.18 | 0.191 |  |  |  |

- i. Time is defined as the time when the respiratory rate measurement was taken and was restricted to between 07:00hr and 20:00hr respectively. - ii. Rainfall (mm) and ambient temperature on the day when RR measurements were taken were retrospectively extracted from the meteorological department in Basse Santa Su town and linked to the RR dataset.
iii. Stunting was defined as the height/length for age z-score <-2 (HAZ/LAZ < -2), wasting was defined as the weight for height z-score < -2 (WHZ < -2) and underweight was defined as the weight for age < -2 (WAZ < -2).

### Intra-observer variability in respiratory rate measurement

A total of 2,311 paired measurements were used in the reliability assessment. 1,159 (50.2%) of the paired measurements were from female subjects and 460 (19.9%) were from children with signs of illness. Overall, the ICC for RR among all children with paired measurements (age 1 to 13 years) was 0.93 (95%CI: 0.93-0.94) whilst the ICC for children without signs of illness was 0.84 (95%CI: 0.83-0.86). Among children aged less than 5 years, the ICC was 0.94 (95%CI: 0.92-0.96) while the estimate for children above 5 years and <14 years was 0.83 (95%CI: 0.82-0.84). The Bland-Altman plots of intra-observer reliability in RR measurement are presented in Figure 3A and Figure 3B. The mean difference between paired RR measurements was small in both children with (1.49 breaths/minute [95%CI: −0.74 to 3.71]) and without (1.04 breaths/minute [95%CI: −3.94 to 6.02]) signs of illness (Figure 3A and Figure 3B). However, the variability between the paired RR measurements among sick children (SD=2.54) was higher than children without (SD=1.13) signs of illness (p<0.001).

**Figure 3:**
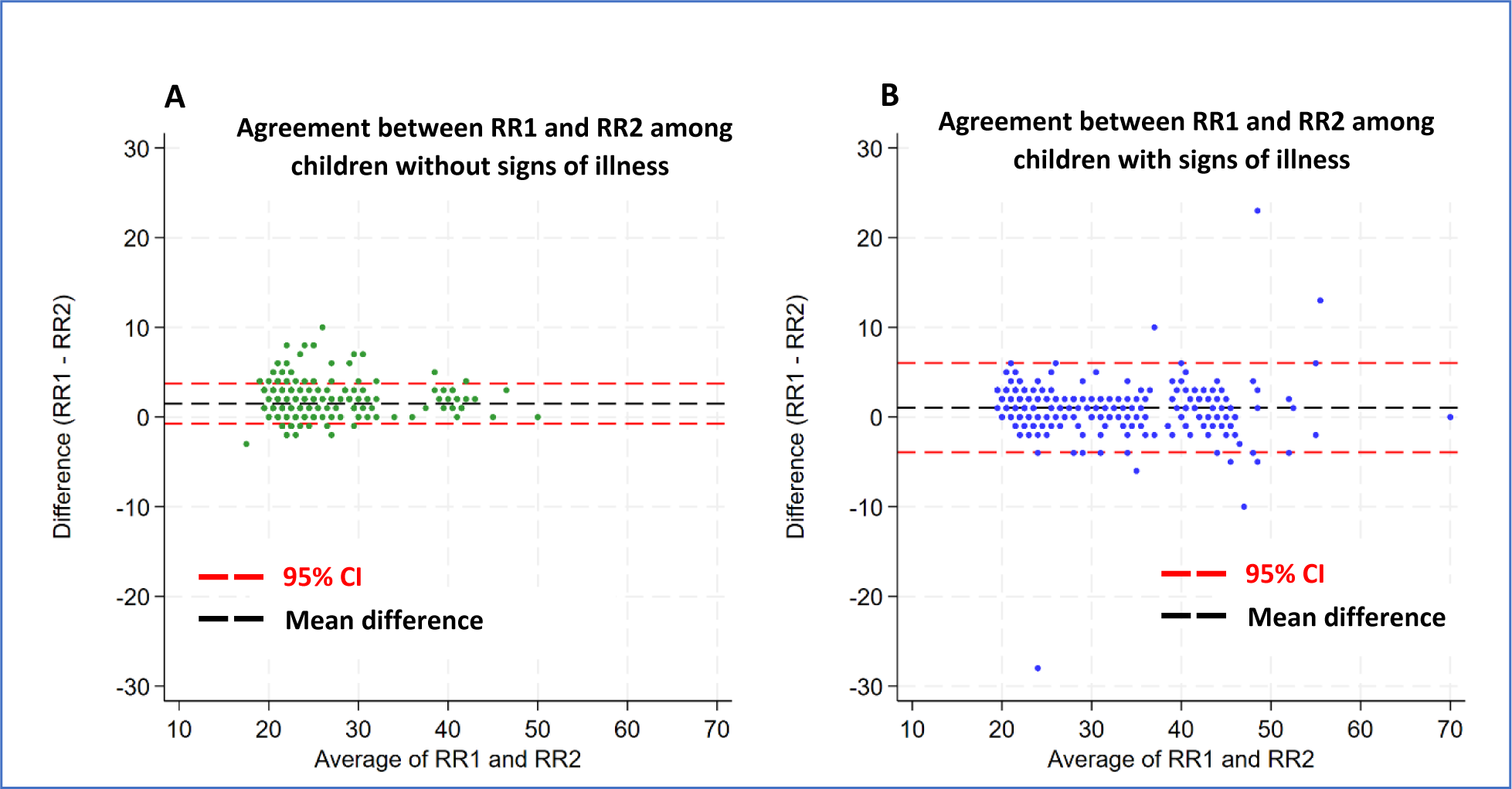
Bland-Altman plots for RR measurements. Panel A displays data from children without signs of illness whilst panel B displays data from children with signs of illness. The black dash line represents the level of bias while the red dash lines display the 95% confidence intervals. RR1 is the first RR measurement while RR2 is the second RR measurement taken in that order. The mean difference in RR measurements was 1.49 breaths/minute (95%CI: −0.74 to 3.71) for children without signs of illness and 1.04 breaths/minute (95%CI: −3.94 to 6.02) for children with signs of illness.

## Discussion

The WHO definition of tachypnoea does not address in any detail children above 5 years of age (29). To construct reference charts for children aged 1 to 13 years we used a longitudinal dataset of RR measurements derived from a large cohort of rural Gambian children enrolled in a trial investigating the impact of improved housing on the incidence of malaria (20,21). These data were linked to potential environmental exposures, demographic factors and three cross-sectional surveys on participant’s nutritional status. RR measurements were done to monitor pneumonia, a potential serious adverse effect, since there was concern that the housing modifications in the intervention arm would increase respiratory disease – however, this was shown not to be the case (20,21). The constructed RR-for-age reference chart among children without signs of illness decreased with increasing age and is consistent with previous findings (17–19).

On average, stunting was associated with a small increase in RR consistent with a recent multicounty pneumonia trial (30) in which stunting was associated with a RR of 1 breath/minute higher at admission to hospital. Whereas the multicounty trial assessed children presenting with signs of pneumonia (30), our study examined RR measurements among mainly healthy children in the community. Stunted children have been reported to have lower ventilatory reserve than children with normal height for age (31). Stunting has been associated with reduced lung function resulting from the impaired lung development (32–34) while chronic undernutrition predisposes children to severe illnesses such as pneumonia that can potentially lead to long-term effects on lung development (33–36). Therefore, the relatively higher RRs observed among stunted children may be compensatory for inadequate ventilation. Our results, however, contrast with a previous study conducted in The Gambia that documented a relatively lower RR measurements among stunted children with signs of pneumonia (37) thus highlighting the need for further research to confirm these findings.

Higher ambient temperature was associated with increased RR measurements consistent with previous studies in both humans and animal models (38). The higher RR observed at higher ambient temperatures may be a homeostatic mechanism by which the body maintains its normal temperature (38). On average, RR measurements taken in the afternoon were higher compared to those taken in the morning or in the evening. Higher RR in the afternoon may be a result of the cumulative increase in the child’s activity during the day. Increased activity (exercise) has been associated with an increase in oxygen requirements leading to an increase in RR (39). Another potential explanation is that ambient temperature increases towards the afternoon and may thus confound the time when measurements were taken. However, the precise ambient temperature when RR measurements were taken was not available for these analyses.

Our study has important strengths. First, there are few studies that provide a comprehensive assessment of the factors associated with RR using a large cohort of children who were actively followed up to document their clinical disease status, nutritional status and other potential environmental risk factors. Second, the RR measurements were taken from children drawn from randomly selected households in the community and therefore the findings are generalizable to the community. In contrast, most RR studies have relied on cross-sectional studies where data are collected in hospital or daycare centers and therefore difficult to generalize to the community. Third, although the repeat RR measurements were only taken among children whose initial measurements were considered raised for age, the intra-observer reliability index (ICC) in both the overall and subgroup analyses were >0.8 suggesting good to excellent reliability (40). In addition, we did not find any systematic differences between the repeat measurement as shown in the Bland Altman plot (Figure 3A and Figure 3B). The variability between the RR repeat measurements was higher among children with signs of illness than among children without signs of illness (p<0.001), highlighting a potential limitation of utilizing a single RR measurement for the classification of danger signs associated with respiratory illness. A limitation to our study is that data were collected from a single site in rural Gambia, a country situated less than 60m above sea-level with high seasonal temperatures. Therefore, the interpretation of our results may be limited to areas with similar geographical and climatic characteristics. However, our study responds to the call for more data from various geographical regions to guide policy development on diagnosis of respiratory illness (7,17).

In conclusion, the current WHO tachypnea guidelines do not address in any detail children above five years of age (29). We constructed centile charts using data collected from a low altitude area in The Gambia and propose a cutoff of >26 breaths/minute for raised RR among children >5 years and below 13 years of age. We demonstrate that age, time of data collection, nutritional status and ambient temperature were associated with RR. However, the small change in RR associated with nutritional status, time of day when RR measurements were taken and ambient temperature does not warrant a change in the current WHO tachypnea guidelines for children > one year and <5 years of age. The finding that RR differences between paired repeat measurements were more variable among children with signs of illness raises concerns over the reliability of a single RR measurements among children with signs of illness - a population in need of accurate diagnosis to enable targeted interventions with lifesaving treatment.

## Data Availability

Data sharing Access to clinical data requires a formal application to the Scientific Coordinating Committee of the Medical Research Council Unit The Gambia (MRCG) and the Joint Gambian Government’s and MRCG Ethics Committee in The Gambia at https://www.mrc.gm/scientific-coordinating-committee/

## Ethical Approval

Ethical approval for the study was granted by The Gambia Government/Medical Research Council Joint Ethics Committee (reference: SCC 1390v3) and the School of Biological and Biomedical Sciences Ethics Committee, Durham University, Durham, UK (reference: SBBS/EC/ 1401/RooPfs). Written informed consent was obtained from the head of the household and/or the parent or guardian of all participating children. The study was conducted in accordance with the International Conference on Harmonization Tripartite Guideline for Good Clinical Practice and the Declaration of Helsinki.

## Data sharing

Access to clinical data requires a formal application to the Scientific Coordinating Committee of the Medical Research Council Unit The Gambia (MRCG) and the Joint Gambian Government’s and MRCG Ethics Committee in The Gambia at https://www.mrc.gm/scientific-coordinating-committee/

## Acknowledgments

The study was funded by the MRC-DfID-Wellcome Trust (Global Health Trials; MR/M007383/1). SWL is supported by the Global Challenges Research Fund’s BOVA Network (Global Challenges Research Fund (BB/R00532X/1) supported by the Biotechnology and Biological Sciences Research Council and Medical Research Council). JB received support from the UK Medical Research Council (MRC) and the UK Foreign, Commonwealth and Development Office (FCDO) under the MRC/FCDO Concordat agreement and is also part of the EDCTP2 programme supported by the European Union (Grant Ref: MR/R010161/1). We are grateful for the support of the villagers, the village health workers, the regional health team, and staff in the health clinics. We also thank the MRCG at the London School of Hygiene and Tropical Medicine for hosting this study in Basse Field Station and their technical and logistical support, especially the nurse field assistants and their supervisors; and the data supervisor, manager, and data entry staff. We are also grateful to members of the Trial Steering Committee and the Data Safety and Monitoring Board.

## Authors Contributions

**Conceptualization and design:** PM, JG, MP, UD’, SL, JB

**Performed the experiments:** MP, UD’, SL, JB

**Data curation:** PM, SA, JG, JB

**Funding acquisition:** MP, SL

**Project administration and investigation:** MP, SL

**Formal data analysis:** PM, SA

**Writing-original draft:** PM, SA

**Writing-review and editing:** PM, SA, JG, MP, LT, UD’, SC, SL, JB,

**All authors have read, and confirm that they meet, ICMJE criteria for authorship.**

---

**Supplementary Figure 1:**
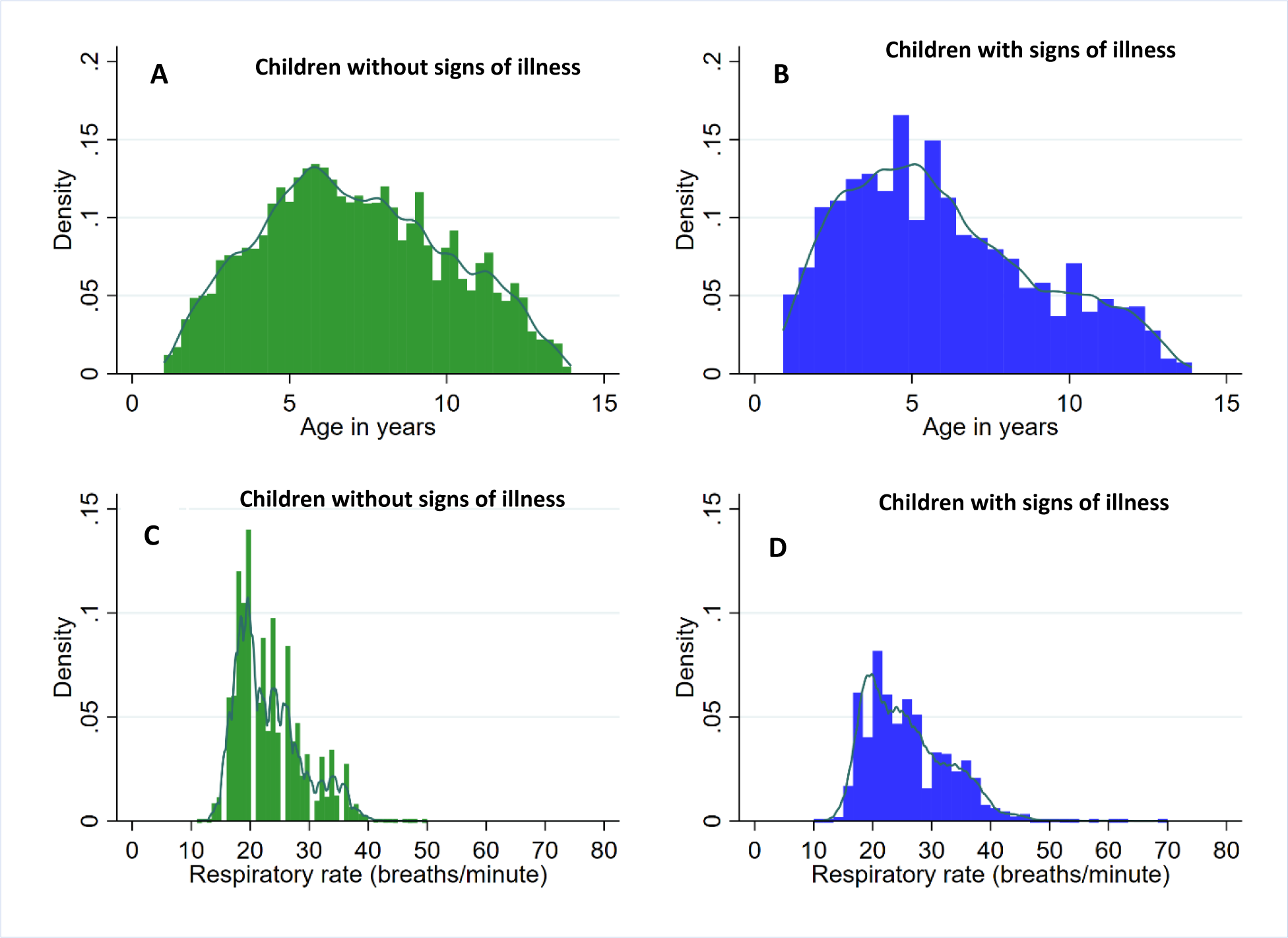
Histograms of the distribution of study participants by age and respiratory rate measurements. Panels A and B show the distribution of age among children without and children with signs of illness respectively and panels C and D show the distribution of respiratory rate among children without and children with signs of illness respectively.

**Supplementary Table 1:** Summary of children with signs of illness, their corresponding observations and median respiratory rate segregated by age.

|  | Children with signs of illness |  |  | Children without signs of illness |  |  |
| --- | --- | --- | --- | --- | --- | --- |
| Age in Years | Number of observations (%) | Number of children | Median RR (IQR) | Observations | Number of Children | Median (IQR) |
| 1 to <1.5 | 60 (13) | 16 | 35 (32 - 39) | 410 | 20 | 32 (28 - 35) |
| 1.5 to <2.5 | 407 (13) | 77 | 32 (29 - 36) | 2609 | 89 | 32 (28 - 35) |
| 2.5 to <3.5 | 492 (10) | 113 | 30 (27 - 34) | 4308 | 142 | 30 (26 - 34) |
| 3.5 to <4.5 | 514 (9) | 136 | 28 (25 - 34) | 5273 | 177 | 28 (24 - 33) |
| 4.5 to <5.5 | 566 (7) | 156 | 26 (22 - 30) | 7180 | 218 | 25 (21 - 30) |
| 5.5 to <6.5 | 510 (6) | 153 | 22 (19 - 24) | 7875 | 237 | 21 (19 - 25) |
| 6.5 to <7.5 | 369 (5) | 133 | 20 (19 - 24) | 6753 | 209 | 20 (19 - 24) |
| 7.5 to <8.5 | 315 (5) | 116 | 20 (18 - 24) | 6593 | 193 | 20 (18 - 23) |
| 8.5 to <9.5 | 227 (4) | 91 | 20 (19 - 24) | 5821 | 167 | 20 (18 - 24) |
| 9.5 to <10.5 | 231 (5) | 84 | 20 (18 - 23) | 4600 | 142 | 19 (18 - 22) |
| 10.5 to <11.5 | 176 (4) | 63 | 19 (18 - 21) | 3881 | 118 | 20 (18 - 22) |
| 11.5 to <12.5 | 179 (5) | 66 | 20 (19 - 23) | 3092 | 92 | 19 (18 - 22) |
| 12.5 to <13.5 | 67 (4) | 26 | 20 (18 - 22) | 1478 | 49 | 19 (17 - 22) |

